# Socio-demographic predictors of Insecticide-treated bed net ownership and utilization for protection against malaria by rural community members across five regions of Mainland Tanzania

**DOI:** 10.1101/2025.10.01.25337095

**Authors:** Gervas A. Chacha, Misago D. Seth, Salehe S. Mandai, Daniel A. Petro, Daniel P. Challe, Angelina J. Kisambale, Rule Budodo, Rashid A. Madebe, Ruth B. Mbwambo, Catherine Bakari, Dativa Pereus, Sijenunu Aaron, Daniel Mbwambo, Samuel Lazaro, Celine I. Mandara, Deus S. Ishengoma

## Abstract

**Background:** Despite decades of control efforts by the National Malaria Control Programme, malaria burden in Tanzania remains high, with transmission intensity varying significantly across regions. Insecticide-treated bed nets (ITNs) are among the core interventions recommended by the World Health Organization for malaria prevention and control in endemic areas. In Tanzania, ITNs are distributed through multiple channels to promote their acquisition and use for protection against malaria. However, persistent gaps in ITN ownership and usage raise concerns about equitable access and utilization. This study aimed to evaluate socio-demographic predictors of ITNs ownership and usage among rural community members in five districts from five regions with varying malaria endemicity.

**Methods:** A cross-sectional survey was conducted in selected communities (covering 15 villages) from five districts, one each from five regions of Kagera, Kigoma, Njombe, Ruvuma, and Tanga, from July to August 2023. Community members (including those with malaria symptoms and asymptomatic participants) aged ≥6 months were recruited. Demographic, malaria prevention practices, anthropometric, clinical, parasitological and socio-economic status (SES) data were captured using mixed questionnaires configured and installed on Open Data Kit (ODK) software, which runs on tablets. Socio-demographic predictors associated with ITNs ownership and use were determined using logistic regression analysis. The results were presented as crude (cOR) and adjusted odds ratios (aOR) with 95% confidence intervals (CI) and a p-value <0.05 was considered statistically significant.

**Results:** Among the 10228 enrolled participants, 7939 (77.6%) and 7899 (77.2%) reported owning and using ITNs, respectively. Both ownership and usage varied significantly across districts (p<0.001), with the highest rates reported in Nyasa and the lowest in Kyerwa. Females were more likely than males to own (aOR=1.27, 95% CI:1.12 - 1.45, p<0.001) and use ITNs (aOR=1.27, 95%CI:1.12 - 1.45, p<0.001). Similarly, under-fives had significantly higher odds of ITN ownership (aOR=1.83, 95%CI:1.56 - 2.15, p<0.001) and usage (aOR=2.26, 95%CI:1.62 - 3.15, p<0.001) than school-children with reference to adults. Participants from Nyasa - Ruvuma, Ludewa - Njombe, and Muheza - Tanga also exhibited higher odds of ITN ownership and usage compared to those from Buhigwe - Kigoma, with Kyerwa - Kagera as the reference district (p<0.001). The odds of ITN ownership and usage increased significantly with an increase in education levels (p≤0.200) and SES (p≤0.001).

**Conclusion:** ITN ownership and usage were relatively high among rural community members in the five surveyed districts; however, both remained below the 80% target projected for 2023. Higher ownership and usage were observed among females, under-fives, individuals with higher education levels, and those from households with higher SES. Disparities based on sex, age groups, socioeconomic status, education, and occupation should be carefully considered in ITN distribution strategies to ensure equitable access and usage of bed nets across all population groups in Tanzania.

## Background

Malaria burden in Tanzania remains substantially high despite decades of significant control efforts led by the National Malaria Control Programme (NMCP) [1]. The insecticide-treated bed nets (ITNs) are among the core interventions that have been recommended by the World Health Organization (WHO) for malaria control and prevention in endemic countries [2,3]. Since 2004, Tanzania NMCP has been implementing the ITNs distribution programs [4,5] and along with other vector control interventions, such as indoor residual spraying (IRS), larval source management (LSM) and enhanced case management (prompt diagnosis and effective treatment) and chemopreventive therapies contributed to the substantial decline of malaria burden from 18.1% in 2008 to 8.1% in 2022 [6,7].

ITNs stand as the cornerstone of malaria vector control interventions aiming at preventing transmission and reducing malaria-related morbidity and mortality among the risk groups [8,9]. Although other vulnerable populations are yet to be identified and targeted, the ITNs distribution programs have primarily targeted infants and pregnant women as the highly vulnerable and high risk groups to malaria [10,11]. The first-ever nationwide ITN distribution campaign in Tanzania, known as the Tanzania National Voucher Scheme (TNVS), was launched in 2004 and initially provided subsidized ITNs to pregnant women and infants [12,13]. Currently, the related program operates through antenatal care (ANC) and immunization visits aiming to keep up and sustain the ITN access to pregnant women and infants [14]. Alongside TNVS, there were other ITNs mass distribution campaigns of 2009 and 2011 that targeted the under-fives and other vulnerable groups, aiming for universal coverage, respectively [15,16]. In 2013, the NMCP initiated school net Programme (SNP) which operates through primary schools and distributes ITNs annually [15,17,18]. Initially covering three regions in consecutive three years, the SNP was scaled up to seven regions in 2016 and further to over half of the country (14 of 26 regions of Mainland Tanzania) in 2017 [17]. The goal of this extensive school based ITNs distribution strategy was to sustain bed net coverage to target levels and ensure the universal access to and use of ITNs rate of at least one ITN for every two people and reaching coverage of 80% by 2023 and 100% by 2025 [5,19].

Besides the ITNs mass-distribution campaigns, there are ongoing behaviour change and communication (BCC) campaigns in Tanzania [20]. The initiatives led by NMCP and its partners utilize various media channels including radios, televisions, posters, flyers, leaflets and billboards, as well as engaging schools, communities, private and public health workers to raise public awareness, encourage acquisition and adoption of ITNs and other interventions to combat malaria [21–23]. BCC campaigns are being implemented in large scale to enhance the use of malaria interventions and ensure their effectiveness while promoting broader improvement in public knowledge, attitude, perception and practices related to malaria control and prevention [23,24]. These campaigns involve the use of communication messages featuring various slogans such as “Malaria haikubaliki” (Malaria is not acceptable) and “Ziro malaria inaanza na mimi” (Zero malaria starts with me) [23,25]. They advocate for malaria risk awareness among pregnant women, infants and under-fives, promoting acquisition and consistent use of ITNs to reduce the risk of contracting malaria. They also emphasize the importance of seeking malaria testing in nearby health facilities before using antimalarial drugs and ensuring proper adherence to treatment and preventing parasite resurgence.

Despite extensive efforts, several challenges compromise the effectiveness of ITNs, including the widespread insecticide resistance among mosquito vectors and reduced net durability and bio-efficacy over time [26,27]. Over the past two decades, Tanzania has reported insecticide resistance to multiple classes of chemicals used in vector control [27,28], leading the country to change the types of insecticides used in ITNs and other interventions such as IRS [5,29]. Several studies have also reported reduced ITNs efficacy falling below WHO recommended standards, with net lifespan limited to two years or less due to insecticidal decay or compromised physical integrity [30,31]. Thus, NMCP should consider adopting innovative ITNs with enhanced bio-efficacy in its procurement and replacement strategies to sustain effective malaria vector control and ensure compliance with the recommended standard for net integrity and lifespan. Operational and behavioural gaps also pose significant barriers to the sustainable and universal adoption of ITNs [19]. These include inadequate distribution channels particularly in rural areas, availability of untreated and unaffordable bed nets in private sectors, and insufficient knowledge and misconceptions that contribute to the misuse of ITNs [32].

In response to heterogeneous transmission patterns and varying malaria burdens across the country, the Tanzania NMCP shifted from “one size fits all” approach to more tailored and stratified malaria control strategies [5,33]. Interventions are now adopted and deployed to sub-regions based on area-specific malaria burden, aiming to lower the disease burden in the high and moderate transmission strata and to achieve pre-elimination or elimination in the low and very low transmission areas, respectively [34,35]. Estimation of ITN access and quantification for programmatic planning are currently done at council level, primarily using household survey data such as Malaria Indicator Survey or Demographic and Health Survey [1,36,37]. However, these surveys are conducted after every three to five years, and availability of data is often delayed due to the time required for fieldworks and analysis, as results are not well-suited for timely, annual monitoring of ITN access. This study aimed to evaluate the socio-demographics determinants of ITNs ownership and usage for malaria prevention among individuals from rural communities across five regions of Tanzania Mainland. The findings will support NMCP and other key malaria control partners to identify and target other vulnerable populations, addressing the socio-demographic gaps in ITNs distributions and strengthening ongoing malaria control interventions towards elimination goal.

## Methods

### Study design and sites

This study utilized data collected from the community-based cross-sectional surveys (CSS) conducted in five regions of Tanzania mainland with different malaria transmission intensity as previously reported [38,39]. Data were collected by the project on Molecular Surveillance of Malaria in Tanzania (MSMT) which involved both health facilities and community surveys. The regions involved in CSS include two each from high (Tanga and Ruvuma) and moderate (Kagera and Kigoma), and one region (Njombe) from low transmission strata. In each region, one district and varying numbers of villages were selected. In Tanga region, the survey covered three villages (Magoda, Mamboleo and Mpapayu) from Muheza district [40]. In Kagera region, five villages were covered (Kitwechenkura, Nyakabwera, Rubuga, Kitoma and Ruko) all from Kyerwa district as previously described [39], while in Kigoma, the surveys were done in two villages (Nyankoronko and Kigege) both from Buhigwe district. In Ruvuma, the study was conducted in four villages (Lundo, Lipingo, Ngindo and Chiulu) from Nyasa district and in Njombe region one village (Kipangala) from Ludewa district was covered (**Figure 1**). The majority of individuals from these villages are served by dispensaries that have been involved in the longitudinal component of the project since 2022 aiming to monitor parasite populations and malaria patterns in these regions, as described elsewhere [38,41,42]

**Figure 1:**
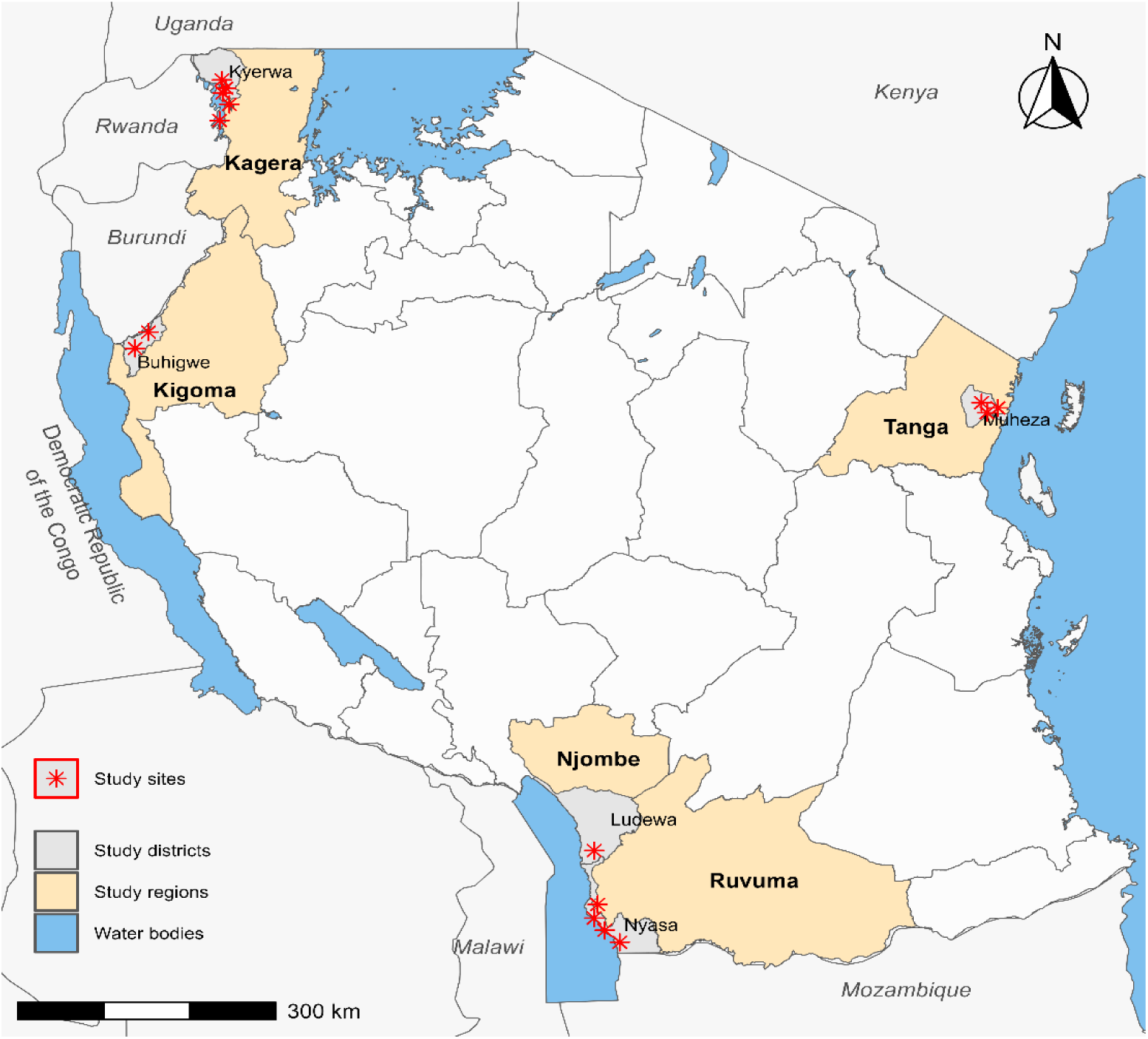
Map of Tanzania showing the regions, districts and study sites/villages that were covered in the community-based cross-sectional surveys in 2023.

### Study population and recruitment process

Data used in this study were collected in the community surveys which aimed to recruit at least 30% all individuals aged ≥6 months from the registered households residing within study villages. In the target communities, household registration was done earlier through the census surveys done by the MSMT project team before the beginning of CSS, as described earlier [38,39]. In brief, the CSS inclusion criteria included individuals aged six months and above, residing in the registered household within the study villages and providing an informed consent. Individuals from villages whose households were not registered and those who declined to provide informed consents were not recruited in this study. All individuals from registered households within the study villages were informed about CSS and invited to participate willingly in the surveys as described recently [38–40].

### Data collection procedures

This study utilized data collected in both census and community cross-sectional surveys as previously described [38,39]. In summary, before CSS, census surveys were conducted to collect demographic data, register households, obtain household socio-economic status (SES) as well as environment, together with land use and geographic information system data. It also involved enumeration of the community members and their households and providing them with unique identification numbers (IDs). During CSS, all participants were identified based on the IDs provided previously in the census survey and data collection proceeded as described earlier [38,39]. In summary, the data collection involved verifying participants’ identities, assigning them study-specific IDs for CSS and issuing the registration cards. Subsequently, all participants were invited to provide consent or assent (for participants aged 7 – 17 years), followed by interviews on socio-demographics and malaria prevention practices. Afterwards, participants were directed to next sections for anthropometric measurements, laboratory screening for malaria parasites using rapid diagnostic tests and blood sample collection onto blood films (thin and thick) and dried blood spots on Whatman filter paper for further laboratory analysis. Lastly, participants were assessed with the study clinicians to collect data on history of illness and any treatment undergone two weeks before surveys, this involved physical and clinical diagnosis, as well as treatment in case of malaria positive cases or any other illnesses, which were managed accordingly [38].

### Data management and analysis

The data were collected using questionnaires prepared and configured in Open Data Kit (ODK) software installed in tablets. Collected data were instantly transferred to the central server at the National Institute for Medical Research (NIMR), Dar es salaam for integration and quality checks. Data discrepancy and unexpected values were highlighted for rejection or prompt correction from the field team. Additional data cleaning was done in Excel and later transferred to STATA version 13 (STATA Corp Inc., TX, USA) for final cleaning and analysis.

Descriptive analysis was performed to provide the baseline information and demographic features of the study populations. The relationships between categorical variables were assessed using chi-square tests. The results were presented in texts, tables and figures. Multilevel logistic regression was used to assess the association between ITNs ownership and utilization and other covariates such as age groups, sex, household size, education levels, and geographic locations. Variables with p<0.25 in the univariate analysis were fitted into multivariate models. Hierarchical model-building strategies were used for further analysis whereby the first model was made by adjusting for individual-level variables such as sex, age group, and the use of bed nets the night before the survey. In the second the analysis, both individual and household characteristics such as household size, household wealth index, and the type of houses inhabited (focusing on the type of windows, walls and presence/absence of eaves) were included. Principal component analysis (PCA) was used to determine the SES of the households based on the assets owned by each family, using the data collected during the census survey to compute the wealth index of the family as previously described [39]. The variables included in the PCA were household possessions and assets such as the occupation of head of households, number of rooms in the house, ownership of items such as mobile phones, motorcycles, bicycles, and domestic animals such as cattle, goats, chickens and pigs. Other assets were land owned by the family and the number of acres cultivated, the source of drinking water, lighting and cooking energy, and the type of toilet. The scores of the first component with eigenvalues >1 were used to create the household wealth index/SES (categorized as high, moderate and low). The association between variables was reported as crude (cOR) or adjusted odds ratios (aOR) with 95% confidence intervals (CIs), and p-value ≤0.05 was considered statistically significant.

## Results

### Baseline characteristics of the study participants

This community-based cross-sectional study was conducted from July to August 2023, involving 10228 individuals from rural communities (with or without symptoms of malaria (across five districts in five regions of Mainland Tanzania. The overall median age of participants was 14 years (Interquartile range (IQR) 7 – 38) and the majority (60.3%, n = 6163) were females, while the remaining (39.7%, n = 4065) were males. Among all participants, 47.9% (n=4899) were adults aged 15 years and older, while 35.6% (n=3640) were school children (aged 5 - <15 years) and the rest (16.5%, n=1689) were under-fives. The distribution of participants varied across the five districts, with Kyerwa accounting for 43.6% (n=4454) of all participants and Nyasa contributing 24.0% (n=2455). The remaining three districts each contributed less than 20.0% of all participants; Buhigwe with 14.2%, (n = 1453), Muheza with 12.3% (n = 1255) and Ludewa had 6.0% (n=611). Of all participants, 20.6% (n = 2102) reported a history of fever in the past 48 hours prior to the survey and 2.3% (n = 238) had fever at presentation. A high proportion of the participants (37.0%, n = 2411) had completed primary education and the main occupation was farming (39.8%, n = 2565). Most of the recruited participants (40.9%, n = 3953) were from households with higher SES while 34.2% (n = 3302) were from households with moderate SES, and 24.9% (n = 2409) were from households with low SES (**Table 1).**

**Table 1:** Baseline characteristics of study participants.

| Characteristics | Ludewa | Buhigwe | Muheza | Kyerwa | Nyasa | Total |
| --- | --- | --- | --- | --- | --- | --- |
| Enrolled, n (%) | 611 (6.0) | 1453 (14.2) | 1255 (12.3) | 4454 (43.6) | 2455 (24.0) | 10228 |
| Sex, n (%) |  |  |  |  |  |  |
| Female | 362 (59.3) | 914 (62.9) | 720 (57.4) | 2641 (59.3) | 1526 (62.2) | 6163 (60.3) |
| Male | 249 (40.8) | 539 (37.1) | 535 (42.6) | 1813 (40.7) | 929 (37.8) | 4065 (39.7) |
| Age in years, Median (IQR) | 14 (7 – 43) | 12 (7 – 26) | 14 (8 – 39) | 14 (7 – 36) | 18 (9 – 45) | 14 (7 – 38) |
| Age groups, n (%) |  |  |  |  |  |  |
| <5 years | 96 (15.7) | 260 (17.9) | 160 (12.7) | 835 (18.8) | 338 (13.8) | 1689 (16.5) |
| 5 - <15 years | 219 (35.8) | 672 (46.2) | 493 (39.3) | 1471 (33.0) | 785 (32.0) | 3640 (35.6) |
| 15+ years | 296 (48.5) | 521 (35.9) | 602 (48.0) | 2148 (48.2) | 1302 (54.3) | 4899 (47.9) |
| History of fever past 48 hours, n (%) |  |  |  |  |  |  |
| Yes | 72 (11.8) | 338 (23.3) | 246 (19.6) | 1342 (30.1) | 104 (4.2) | 2102 (20.6) |
| No | 539 (88.2) | 1115 (76.7) | 1009 (80.4) | 3112 (69.9) | 2351 (95.8) | 8126 (79.4) |
| Fever at presentation ($\geq 37.5^{\circ}\text{C}$), n (%) | | | | | | |
| Yes | 5 (0.8) | 25 (1.7) | 21 (1.7) | 136 (3.1) | 51 (2.1) | 238 (2.3) |
| No | 606 (99.2) | 1428 (98.3) | 1234 (98.3) | 4318 (96.9) | 2404 (97.9) | 9990 (97.7) |
| Education level, n (%) |  |  |  |  |  |  |
| None | 12 (3.0) | 92 (9.3) | 40 (5.9) | 566 (20.5) | 75 (4.5) | 785 (12.0) |
| Incomplete primary | 107 (26.3) | 362 (36.4) | 225 (32.9) | 827 (30.0) | 507 (30.2) | 2028 (31.1) |
| Completed primary | 199 (48.9) | 266 (26.7) | 263 (38.5) | 905 (32.8) | 778 (46.3) | 2411 (37.0) |
| Secondary or above | 21 (5.2) | 10 (1.0) | 28 (4.1) | 75 (2.7) | 87 (5.2) | 221 (3.4) |
| Studying | 68 (16.7) | 265 (26.6) | 128 (18.7) | 386 (14.0) | 232 (13.8) | 1079 (16.5) |
| Occupation, n (%) |  |  |  |  |  |  |
| Farmer | 232 (41.9) | 60 (16.0) | 6 (6.1) | 1634 (41.5) | 633 (42.7) | 2565 (39.8) |
| Students | 155(28.0) | 137 (36.6) | 32 (32.7) | 1061 (26.9) | 469 (31.7) | 1854 (28.8) |
| Children | 147 (26.5) | 160 (42.8) | 52 (53.1) | 1180 (30.0) | 305 (20.6) | 1844 (28.6) |
| Others | 20 (3.6) | 17 (4.6) | 8 (8.2) | 64 (1.6) | 74 (5.0) | 183 (2.8) |
| Socio-economic status, n (%) |  |  |  |  |  |  |
| Higher | 253 (42.6) | 598 (45.2) | 833 (67.7) | 1102 (26.7) | 1167 (48.9) | 3953 (40.9) |
| Moderate | 287 (48.3) | 439 (33.2) | 274 (22.2) | 1516 (36.7) | 786 (32.9) | 3302 (34.2) |
| Low | 54 (9.1) | 286 (21.6) | 124 (10.1) | 1509 (36.6) | 436 (18.2) | 2409 (24.9) |

**n= Number of observed participants; IQR= Inter-quantile range, SES= Socio-economic status; °C= degree Celsius**

### Insecticide treated bed net ownership among study participants

Among 10228 participants enrolled, 77.6% (n = 7939) reported owning insecticide-treated bed nets. The ownership rates varied significantly across districts (p<0.001), with the highest rate in Nyasa (92.1%, n=2262/2455) and the lowest rate in Kyerwa (64.4%, n=2867/4454). Females had higher bed net ownership compared to males in all districts with significant variations in Muheza (p<0.001) and Kyerwa district (p=0.006). Bed net ownership varied significantly with age groups in all five districts (p≤0.006), under-fives demonstrated higher ownership in all districts except in Muheza where school-children (aged 5 - <15 years) surpassed them. The least bed net ownership was observed among adults (aged 15+ years) in Buhigwe, Muheza and Nyasa districts and school-children in Ludewa and Kyerwa districts (**Figure 2**). Participants who reported a history of fever (in the past two days) had lower bed net ownership than those with no history of fever in all districts, except in Nyasa, with significant difference observed in Muheza district (p=0.004). However, bed net ownership remained statistically similar for individuals with/without fever at presentation. In Buhigwe, Muheza, Kyerwa and Nyasa districts individuals with secondary education or above had higher bed net ownership while those with no formal education had the lowest ownership (p≤0.005). In Ludewa district, participants with secondary education or above had the least ownership and higher rates (>90%) were observed among those with no education, studying or those who completed primary education, but their differences were not statistically significant (p=0.662). Individuals working in other sectors than farms, students and children reported higher bed net ownership in all districts except in Nyasa, with significant statistical differences (p≤0.002) observed only in Kyerwa and Nyasa districts. Individuals from households with higher SES demonstrated higher bed net ownership in all five districts. The least ownership was observed among individuals from households with lower SES except in Ludewa district where those from moderate SES had the lowest ownership (p=0.056). (**Table 2a**).

**Table 2a:** Bed net ownership among participants in five surveyed districts.

| Variable | Ludewa | Buhigwe | Muheza | Kyerwa | Nyasa | Total |
| --- | --- | --- | --- | --- | --- | --- |
| Enrolled, N | 611 | 1453 | 1255 | 4454 | 2455 | 10228 |
| Bed net ownership, n (%) | 555 (90.8) | 1101 (75.8) | 1154 (92.0) | 2867 (64.4) | 2262 (92.1) | 7939 (77.6) |
| Sex, n (%) |  |  |  |  |  |  |
| Female | 330 (91.2) | 694 (75.9) | 683 (94.9) | 1743 (66.0) | 1414 (92.7) | 4864 (78.9) |
| Male | 225 (90.4) | 407 (75.5) | 471 (88.0) | 1124 (62.0) | 848 (91.3) | 3075 (75.7) |
| p-value | 0.737 | 0.857 | <0.001 | 0.006 | 0.218 | <0.001 |
| Age groups, n (%) |  |  |  |  |  |  |
| <5 years | 95 (99.0) | 215 (82.7) | 148 (92.5) | 618 (74.0) | 325 (96.2) | 1401 (83.0) |
| 5 - <15 years | 192 (87.7) | 522 (77.7) | 477 (96.8) | 819 (55.9) | 739 (94.1) | 2749 (75.5) |
| 15+ years | 268 (90.5) | 364 (69.9) | 529 (87.9) | 1430 (66.6) | 1198 (89.9) | 3789 (77.3) |
| p-value | 0.006 | <0.001 | <0.001 | <0.001 | <0.001 | <0.001 |
| History of fever past 48 hours, n (%) |  |  |  |  |  |  |
| Yes | 64 (88.9) | 245 (72.5) | 216 (87.8) | 851 (63.4) | 95 (91.4) | 1471 (70.0) |
| No | 491 (91.1) | 856 (76.8) | 938 (93.0) | 2061 (64.8) | 2167 (92.2) | 6468 (79.6) |
| p-value | 0.542 | 0.107 | 0.008 | 0.382 | 0.759 | <0.001 |
| Fever at presentation ($\geq 37.5^{\circ}\text{C}$), n (%) | | | | | | |
| Yes | 5 (100) | 20 (80) | 19 (90.5) | 85 (62.5) | 45 (88.2) | 174 (73.1) |
| No | 550 (90.8) | 1081 (75.7) | 1135 (92) | 2782 (64.4) | 2217 (92.2) | 7765 (77.7) |
| p-value | 0.476 | 0.619 | 0.802 | 0.644 | 0.295 | 0.091 |
| Education level, n (%) |  |  |  |  |  |  |
| None | 11 (91.7) | 53 (57.6) | 33 (82.5) | 337 (59.5) | 60 (80) | 494 (62.9) |
| Incomplete primary | 94 (87.8) | 277 (76.5) | 213 (94.7) | 498 (60.2) | 469 (92.5) | 1551 (76.5) |
| Completed primary | 184 (92.5) | 201 (75.6) | 233 (88.6) | 651 (71.9) | 706 (90.8) | 1975 (81.9) |
| Secondary or above | 18 (85.7) | 10 (100) | 27 (96.4) | 55 (73.3) | 84 (96.6) | 194 (87.8) |
| Studying | 62 (91.2) | 204 (77.0) | 123 (96.1) | 218 (56.5) | 215 (92.7) | 822 (76.2) |
| p-value | 0.662 | 0.001 | 0.005 | <0.001 | 0.002 | <0.001 |
| Occupation, n (%) |  |  |  |  |  |  |
| Farmer | 213 (91.8) | 45 (75.0) | 5 (83.3) | 1108 (67.8) | 575 (90.8) | 1946 (75.9) |
| Students | 137 (88.4) | 108 (78.8) | 29 (90.6) | 604 (56.9) | 446 (95.1) | 1324 (71.4) |
| Children | 137 (93.2) | 125 (78.1) | 47 (90.4) | 780 (66.1) | 294 (96.4) | 1383 (75.0) |
| Others | 19 (95.0) | 14 (82.4) | 8 (100) | 47 (73.4) | 71 (95.6) | 159 (86.9) |
| p-value | 0.431 | 0.906 | 0.747 | <0.001 | 0.002 | <0.001 |
| Socio-economic status, n (%) |  |  |  |  |  |  |
| Higher | 237 (93.7) | 464 (77.6) | 768 (92.2) | 768 (69.7) | 1090 (93.4) | 3327 (77.9) |
| Moderate | 252 (87.8) | 337 (76.8) | 255 (93.1) | 993 (65.5) | 732 (93.1) | 2569 (77.8) |
| Low | 50 (92.6) | 197 (68.9) | 109 (87.9) | 894 (59.2) | 378 (86.7) | 1628 (67.6) |
| p-value | 0.056 | 0.014 | 0.194 | <0.001 | <0.001 | <0.001 |

**Table 2b:** Bed net usage among participants across five selected districts.

| Variable | Ludewa | Buhigwe | Muheza | Kyerwa | Nyasa | Total |
| --- | --- | --- | --- | --- | --- | --- |
| Enrolled, N | 611 | 1453 | 1255 | 4454 | 2455 | 10228 |
| Bed net usage, n (%) | 555 (90.8) | 1094 (75.3) | 1145 (91.2) | 2849 (64.0) | 2256 (91.9) | 7899 (77.2) |
| Sex, n (%) |  |  |  |  |  |  |
| Female | 330 (91.2) | 689 (75.4) | 680 (94.4) | 1734 (65.7) | 1409 (92.3) | 4842 (78.6) |
| Male | 225 (90.4) | 405 (75.5) | 465 (86.9) | 1115 (61.5) | 847 (91.2) | 3057 (75.2) |
| p-value | 0.737 | 0.917 | <0.001 | 0.005 | 0.307 | <0.001 |
| Age groups, n (%) |  |  |  |  |  |  |
| <5 years | 95 (99.0) | 214 (82.3) | 148 (92.5) | 616 (73.7) | 325 (96.2) | 1398 (82.7) |
| 5 - <15 years | 192 (87.7) | 518 (77.1) | 473 (95.9) | 815 (55.4) | 738 (94.0) | 2736 (75.2) |
| 15+ years | 268 (90.5) | 362 (69.5) | 524 (87.0) | 1418 (66.0) | 1193 (89.6) | 3765 (76.9) |
| p-value | 0.006 | <0.001 | <0.001 | <0.001 | <0.001 | <0.001 |
| History of fever past 48 hours, n (%) |  |  |  |  |  |  |
| Yes | 64 (88.9) | 243 (71.9) | 213 (86.6) | 850 (63.4) | 94 (90.4) | 1464 (69.7) |
| No | 491 (91.1) | 851 (76.3) | 932 (92.4) | 1999 (64.2) | 2162 (92.0) | 6435 (79.2) |
| p-value | 0.542 | 0.098 | 0.004 | 0.567 | 0.564 | <0.001 |
| Fever at presentation ($\geq 37.5^{\circ}\text{C}$), n (%) | | | | | | |
| Yes | 5 (100) | 20 (80) | 19 (90.5) | 84 (61.8) | 44 (86.3) | 172 (72.3) |
| No | 550 (90.8) | 1074 (75.2) | 1126 (91.3) | 2765 (64.0) | 2212 (92.0) | 7727 (77.4) |
| p-value | 0.476 | 0.582 | 0.901 | 0.587 | 0.137 | 0.065 |
| Education level, n (%) |  |  |  |  |  |  |
| None | 11 (91.7) | 53 (57.6) | 33 (82.5) | 336 (59.4) | 59 (78.7) | 492 (62.7) |
| Incomplete primary | 94 (87.9) | 274 (75.7) | 209 (92.9) | 491 (59.4) | 468 (92.3) | 1536 (75.7) |
| Completed primary | 184 (92.5) | 200 (75.2) | 231 (87.8) | 645 (71.1) | 702 (90.2) | 1960 (81.3) |
| Secondary or above | 18 (85.7) | 10 (100) | 27 (96.4) | 55 (73.3) | 84 (96.6) | 194 (87.8) |
| Studying | 62 (91.2) | 204 (77.0) | 122 (95.3) | 218 (56.5) | 215 (92.7) | 821 (76.1) |
| p-value | 0.662 | 0.001 | 0.021 | <0.001 | 0.001 | <0.001 |
| Occupation, n (%) |  |  |  |  |  |  |
| Farmer | 213 (91.8) | 45 (75.0) | 5 (83.3) | 1099 (67.3) | 574 (90.7) | 1936 (75.5) |
| Students | 137 (88.4) | 106 (77.4) | 29 (90.6) | 589 (56.4) | 446 (95.1) | 1316 (71.0) |
| Children | 137 (93.2) | 125 (78.1) | 47 (90.4) | 778 (65.9) | 294 (96.4) | 1381 (74.9) |
| Others | 19 (95.0) | 14 (82.4) | 8 (100) | 46 (71.9) | 70 (94.6) | 157 (85.8) |
| p-value | 0.431 | 0.923 | 0.747 | <0.001 | 0.002 | <0.001 |
| Socio-economic status, n (%) |  |  |  |  |  |  |
| Higher | 237 (93.7) | 461 (77.1) | 763 (91.6) | 765 (69.4) | 1086 (93.1) | 3312 (83.8) |
| Moderate | 252 (87.8) | 334 (76.1) | 252 (92.0) | 987 (65.1) | 730 (92.9) | 2555 (77.4) |
| Low | 50 (92.6) | 197 (68.9) | 108 (87.1) | 885 (58.7) | 378 (86.7) | 1618 (67.2) |
| p-value | 0.056 | 0.025 | 0.226 | <0.001 | <0.001 | <0.001 |

**Figure 2:**
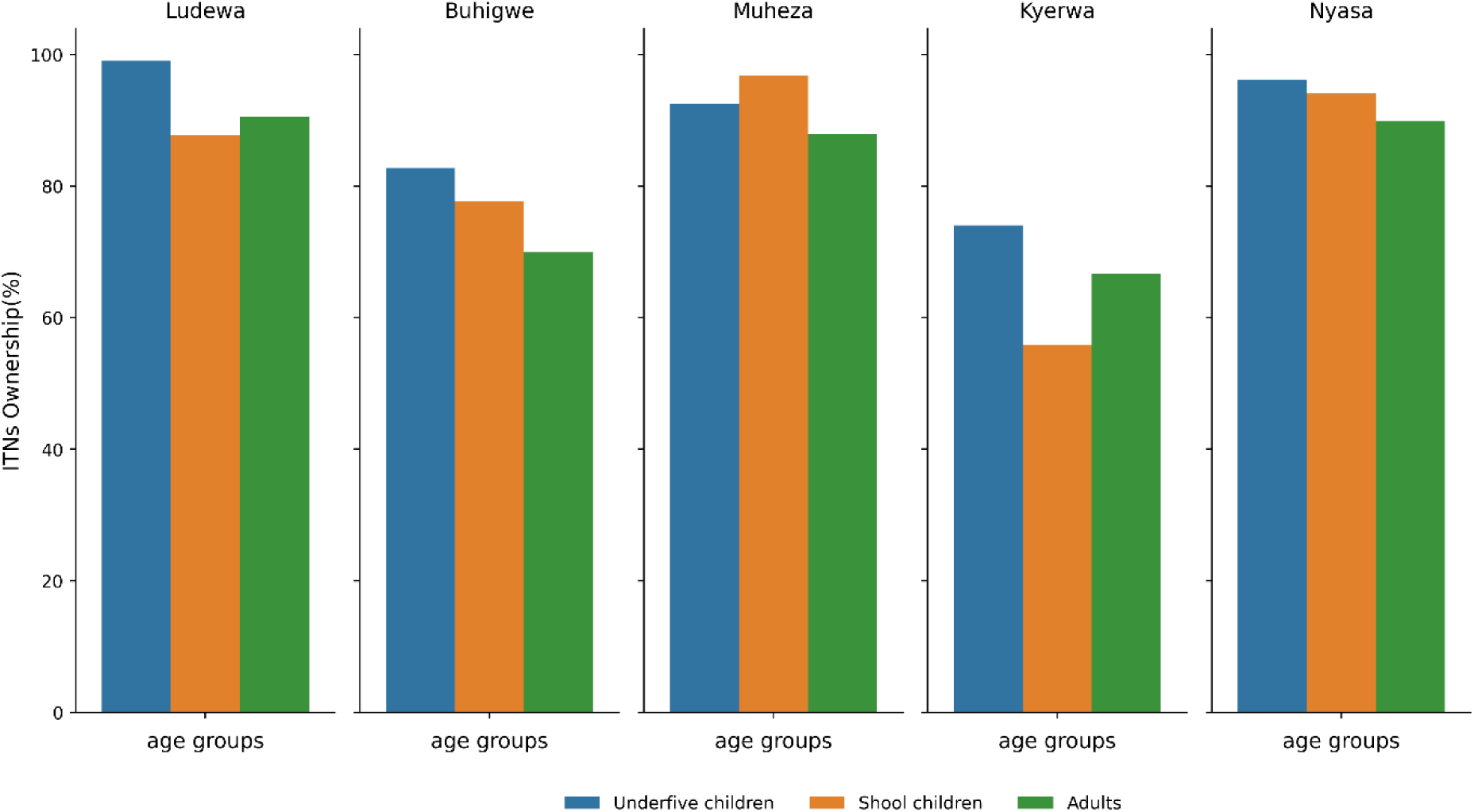
Variation in ITN ownership by age groups among participants in the study districts.

### Insecticide treated bed net usage among study participants

Of the recruited participants in this study, 77.2% (n=7899/10288) reported using insecticide-treated bed nets for protection against mosquito bites. Bed net usage was highest in Nyasa (91.9%, n=2256/2455) and the lowest was in Kyerwa district (64.0%, n=2899/4454) with significant differences observed across districts (p<0.001). Females exhibited higher bed net usage in all districts, except in Buhigwe where bed net usage rate was similar in both sexes (p=0.917). Bed net usage varied significantly with age groups in all five districts (p≤0.006), under-fives showed higher usage in all districts except in Muheza where school-children had higher usage while the lowest usage rates were observed among school-children in Kyerwa and Ludewa as well as adults in Buhigwe, Muheza and Nyasa districts (**Figure 3**). Participants who reported a history of fever in the past two days had lower bed net usage than those with no history of fever, with statistical differences observed in Kyerwa (p=0.004). Overall, no significance differences were seen in bed net usage among individuals with/without fever at presentation (p=0.065). However, higher usage was reported among those who had fever at presentation in Ludewa and Buhigwe districts which was opposite in other districts. Bed net usage varied significantly with education levels (p<0.001). Individuals with secondary education or above had the highest bed net use while those with none or incomplete primary education had the lowest usage in all districts (p≤0.021), except in Ludewa where those who had secondary education or above had the lowest usage, while those completed primary school had the highest usage (p=0.0662). Individuals from other sectors than farming, students or children had higher bed net usage in all districts except in Nyasa where children had the highest usage, while the lowest usage was among students in Ludewa (p=0.431) and Kyerwa (p<0.001) as well as farmers in Buhigwe (p=0.923), Muheza (p=0.747) and Nyasa (p=0.002). Participants from households with higher SES had higher bed net usage except in Nyasa (p=0.226). Lower usage was observed among those from households with lower SES in all districts. Overall, bed net usage varied significantly with SES in Buhigwe, Kyerwa and Nyasa (p≤0.025) (**Table 2b**).

**Figure 3:**
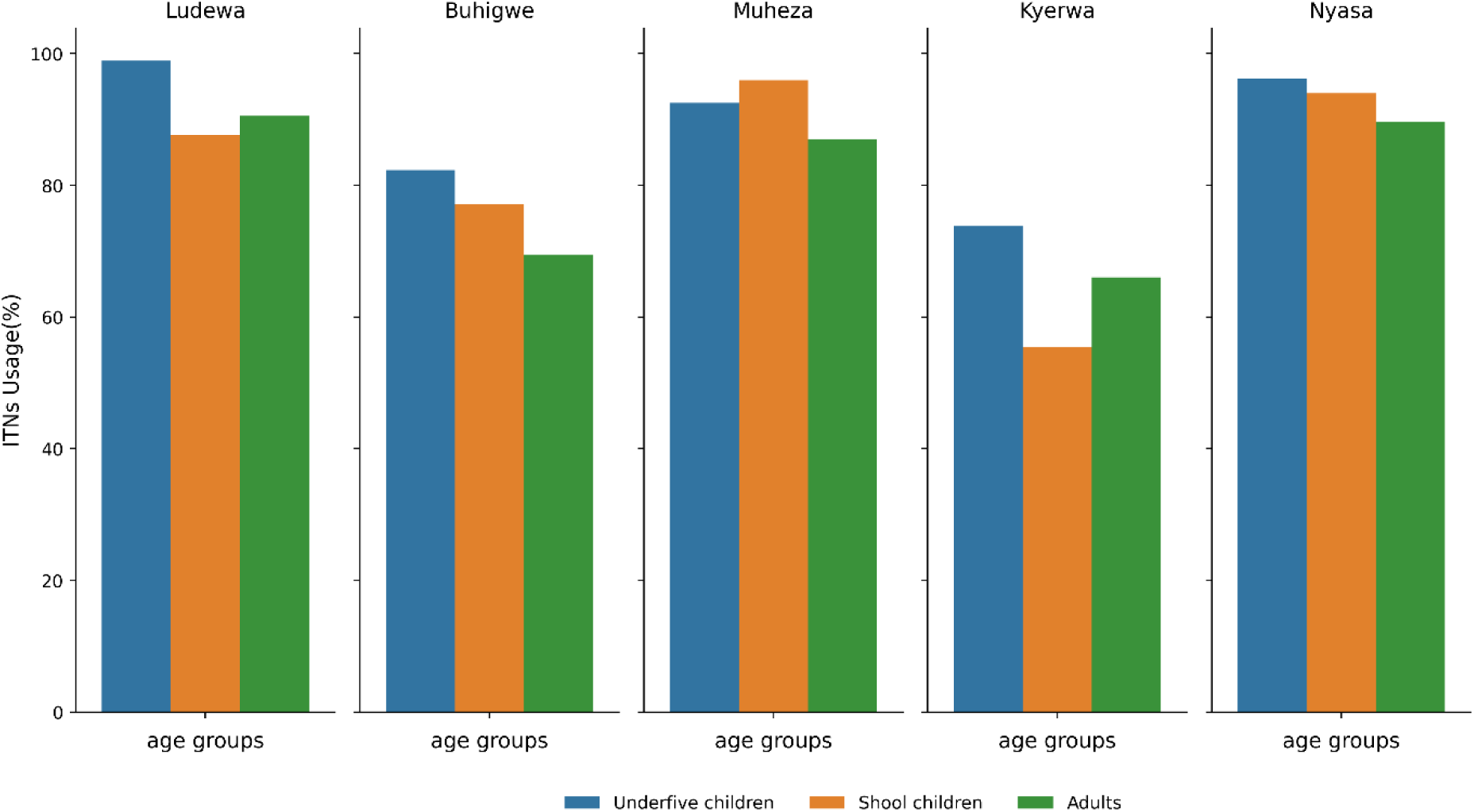
Variations in ITN usage by age groups among participants in the study districts

### Factors influencing ITNs ownership and usage among study participants

Multivariate logistic analysis was done to evaluate factors associated with bed net ownership and usage after adjusting for different confounding variables, and revealed that females had higher odds (aOR=1.27, 95% CI:1.12 - 1.45, p<0.001) of bed net ownership and usage compared to males. Higher odds were observed consistently in under-fives for bed net ownership (aOR=1.83, 95%CI:1.56 - 2.15, p<0.001) and usage (aOR=2.26, 95%CI:1.62 - 3.15, p<0.001) compared to adults. There was no significance difference in ownership and usage of bed nets between school-children and adults. Individuals from Ludewa, Muheza and Nyasa district were more likely (aOR >4.00 with varying 95% CI, p<0.001) to own and use bed nets than those in Kyerwa districts. In Buhigwe, higher odds were also seen in bed net ownership (aOR=1.55, 95% CI:1.31 - 1.85, p<0.001) and usage (aOR=1.57, 95% CI:1.32 - 1.86, p<0.001) in comparison to Kyerwa. Compared to participants who had no formal education, the odds of bed net ownership increased with an increase in the level of education. The odds of bed net ownership and use were higher among participants who were studying (aOR=1.21, 95%CI:0.90 - 1.62, p=0.200), those who had incomplete education (aOR=1.27, 95%CI:1.02 - 1.59, p=0.036), individuals who completed primary education (aOR=1.60, 95%CI:1.33 - 1.94, p<0.001) and those who had secondary education or above (aOR=2.17, 95%CI:1.39 - 3.41, p=0.001). Individuals from households with higher (aOR=1.61, 95%CI:1.36 - 1.90, p<0.001) and moderate SES (aOR=1.31, 95%CI: 1.13 - 1.53, p=0.001) were more likely to own bed net and use them [aOR=1.60, 95%CI:1.36 - 1.89, p<0.001(for higher SES) and aOR=1.31, 95%CI: 1.12 - 1.52, p=0.001(for moderate SES)] compared to individual from households with lower SES (**Table 3**).

**Table 3:**
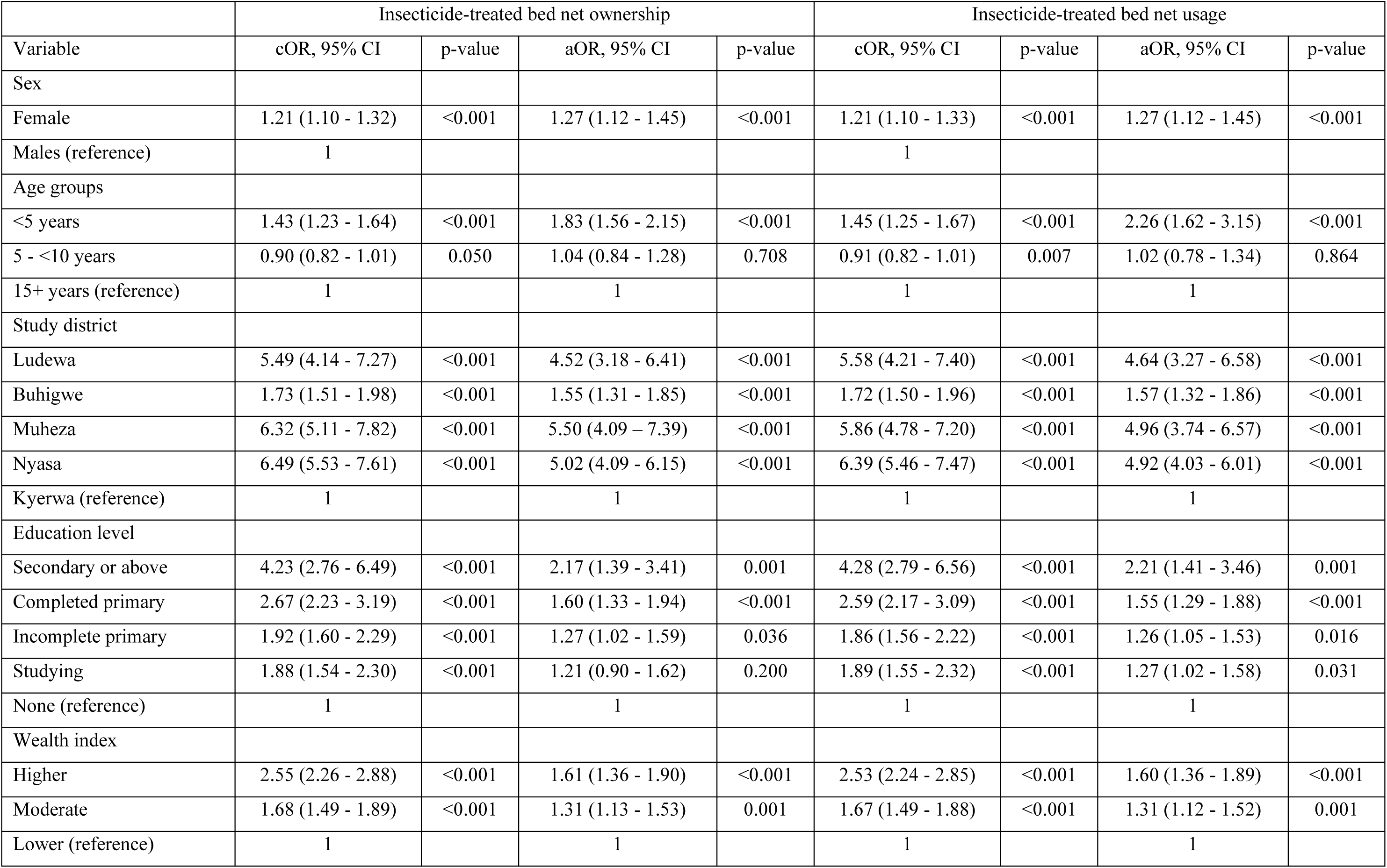
Factors associated with insecticide-treated bed nets ownership and usage.

|  | Insecticide-treated bed net ownership |  |  |  | Insecticide-treated bed net usage |  |  |  |
| --- | --- | --- | --- | --- | --- | --- | --- | --- |
| Variable | cOR, 95% CI | p-value | aOR, 95% CI | p-value | cOR, 95% CI | p-value | aOR, 95% CI | p-value |
| Sex |  |  |  |  |  |  |  |  |
| Female | 1.21 (1.10 - 1.32) | <0.001 | 1.27 (1.12 - 1.45) | <0.001 | 1.21 (1.10 - 1.33) | <0.001 | 1.27 (1.12 - 1.45) | <0.001 |
| Males (reference) | 1 |  |  |  | 1 |  |  |  |
| Age groups |  |  |  |  |  |  |  |  |
| <5 years | 1.43 (1.23 - 1.64) | <0.001 | 1.83 (1.56 - 2.15) | <0.001 | 1.45 (1.25 - 1.67) | <0.001 | 2.26 (1.62 - 3.15) | <0.001 |
| 5 - <10 years | 0.90 (0.82 - 1.01) | 0.050 | 1.04 (0.84 - 1.28) | 0.708 | 0.91 (0.82 - 1.01) | 0.007 | 1.02 (0.78 - 1.34) | 0.864 |
| 15+ years (reference) | 1 |  | 1 |  | 1 |  | 1 |  |
| Study district |  |  |  |  |  |  |  |  |
| Ludewa | 5.49 (4.14 - 7.27) | <0.001 | 4.52 (3.18 - 6.41) | <0.001 | 5.58 (4.21 - 7.40) | <0.001 | 4.64 (3.27 - 6.58) | <0.001 |
| Buhigwe | 1.73 (1.51 - 1.98) | <0.001 | 1.55 (1.31 - 1.85) | <0.001 | 1.72 (1.50 - 1.96) | <0.001 | 1.57 (1.32 - 1.86) | <0.001 |
| Muheza | 6.32 (5.11 - 7.82) | <0.001 | 5.50 (4.09 - 7.39) | <0.001 | 5.86 (4.78 - 7.20) | <0.001 | 4.96 (3.74 - 6.57) | <0.001 |
| Nyasa | 6.49 (5.53 - 7.61) | <0.001 | 5.02 (4.09 - 6.15) | <0.001 | 6.39 (5.46 - 7.47) | <0.001 | 4.92 (4.03 - 6.01) | <0.001 |
| Kyerwa (reference) | 1 |  | 1 |  | 1 |  | 1 |  |
| Education level |  |  |  |  |  |  |  |  |
| Secondary or above | 4.23 (2.76 - 6.49) | <0.001 | 2.17 (1.39 - 3.41) | 0.001 | 4.28 (2.79 - 6.56) | <0.001 | 2.21 (1.41 - 3.46) | 0.001 |
| Completed primary | 2.67 (2.23 - 3.19) | <0.001 | 1.60 (1.33 - 1.94) | <0.001 | 2.59 (2.17 - 3.09) | <0.001 | 1.55 (1.29 - 1.88) | <0.001 |
| Incomplete primary | 1.92 (1.60 - 2.29) | <0.001 | 1.27 (1.02 - 1.59) | 0.036 | 1.86 (1.56 - 2.22) | <0.001 | 1.26 (1.05 - 1.53) | 0.016 |
| Studying | 1.88 (1.54 - 2.30) | <0.001 | 1.21 (0.90 - 1.62) | 0.200 | 1.89 (1.55 - 2.32) | <0.001 | 1.27 (1.02 - 1.58) | 0.031 |
| None (reference) | 1 |  | 1 |  | 1 |  | 1 |  |
| Wealth index |  |  |  |  |  |  |  |  |
| Higher | 2.55 (2.26 - 2.88) | <0.001 | 1.61 (1.36 - 1.90) | <0.001 | 2.53 (2.24 - 2.85) | <0.001 | 1.60 (1.36 - 1.89) | <0.001 |
| Moderate | 1.68 (1.49 - 1.89) | <0.001 | 1.31 (1.13 - 1.53) | 0.001 | 1.67 (1.49 - 1.88) | <0.001 | 1.31 (1.12 - 1.52) | 0.001 |
| Lower (reference) | 1 |  | 1 |  | 1 |  | 1 |  |

## Discussion

Insecticide-treated bed nets are well-designed and stable malaria prevention and control intervention recommended in malaria endemic regions globally. ITNs provide protection against malaria by deterring and killing mosquitoes. ITN distribution programs aim at increasing bed net ownership and sustain access and use to ensure the anticipated impact. Despite increasing distribution efforts, the impact of socio-demographic inequalities on ITNs ownership and usage particularly in rural communities remains unclear. This study was conducted to evaluate socio- demographic determinants of ITNs ownership and utilization for malaria protection among individuals recruited from rural communities in five districts of mainland Tanzania.

In the current study, the overall ITNs ownership and usage were 77.6% and 77.2% respectively, with both varying significantly across all five districts. The study also revealed higher ITNs ownership and usage among females, under-fives, individuals with no history of fever (in the past two days) or those who had no fever at presentation. The ownership and use were also significantly higher among participants with secondary education or above as well as from households with higher SES. The ownership and usage reported in this study were relatively higher compared to the findings reported previously in Tanzania and elsewhere [6,43,44]. The observed high rates are likely attributed by extensive ITNs distribution efforts that have intensively been done by NMCP and its partners, including annual distribution as keep up strategy to sustain high ITNs coverage to ensure universal access and use at both individual and household levels by 2025 [4,5,18,45]. Open marketing has also increased the ITNs retailing channels making them available even in hard-to-reach areas, ultimately leading to increased bed nets access and use [46,47].

There was higher ITNs ownership and usage in females and under-fives across districts. Females reported 78.9 % and 78.6%, and under-fives reported 83.0% and 82.7% in ownership and usage, respectively, and exceeded the average nationwide usage in under-fives (64%) and women (66%) which was reported in Tanzania Malaria Indicator Survey of 2022 and elsewhere [6,48]. In Tanzania, pregnant women and under-fives are given free bed nets during their antenatal care and immunization visits, as a strategy to protect them from malaria during pregnancy and after delivery [49–51]. It also reflects the impact of BCC campaigns fostering positive behavioral changes that have mainly targeted women and under-fives encouraging consistent and correct use of bed nets [22,52]. Among school children, ITNs ownership and use was 75.5% and 75.2%, respectively, aligning with findings reported elsewhere [38]. This consistency is likely due to targeted bed net redistribution programs that identified and recommended for targeting schools as an important channel for sustaining ITNs coverage in Tanzania, following the 2011 national wide mass campaign [15,17,18,53]. Since 2013, several initiatives including SNP have been implemented and scaled-up annually to ensure continued access and use of bed nets [4,54,55]. Despite these efforts, several studies described school-aged children as a group that was highly affected by malaria, which invoked further investigation to disentangle underlying causes of their continued vulnerability [38,56–58]. The lower ITNs ownership and usage in males is probably due to the lack of ongoing ITN distribution initiative targeting this group, particularly adult males as well as those in high risk occupations such as night guards [56,59]. Therefore, as NMCP strives for universal ITNs ownership and use, it is important to assess gender disparities to inform better implementation of future programs. Males also avoid using ITNs due to the misconception and false beliefs leading some to reject free ITNs such as the notion that insecticide used in the bed nets affect fertility including capacity to impair men’s sexuality [19,24,60].

This study also found that individuals with a history fever or fever at presentation had relatively lower ITNs ownership and usage than their counterparts which aligns with findings from studies done in Tanzania and elsewhere, which revealed high malaria susceptibility among non-bed net users [38,39,61]. This is obvious and expected because sleeping under ITNs offers protection against mosquito bites [62–64]. Hence, there is a need for increased sensitization on the benefits of consistent ITN use to achieve intended impacts of distributed bed nets. Additionally, malaria data from the same survey revealed slight variation between ownership and sleeping against malaria protection suggesting a need for thorough investigation on protective efficacy of distributed ITNs [38,65,66].

In this study ITNs ownership and usage increased with an increase in the level of education, with a notable gap between those with no formal education and those with at least some primary education, participants who were still studying or those with much higher levels of education. This is in congruent with findings reported in studies done elsewhere [48,67,68]. This increase could be due to the fact that individuals with primary or higher education are more likely to have learned about malaria prevention in schools and understand better malaria-related messages communicated through leaflets, radio or television [69–71]. Targeted strategies such as deploying health education intervention are needed to reach those with little or no formal education and provide them with comprehensive information on malaria control interventions to enhance knowledge on acquisition and use of ITNs. The higher ITNs ownership and usage among individuals working in non-farming sectors such as business, fishing, and formal employment than those working in farms is consistent with findings reported elsewhere [43,72]. This can largely be attributed to relatively greater financial capacity that enables them to afford the cost to purchase ITNs from commercial outlets which are neither subsidized nor freely distributed [43,73]. Additionally, employees particularly those in formal sectors such as education, health and community leadership are better informed about malaria prevention through their education and qualified roles. They are actively involved in prevention initiatives to raise community awareness and promote access to and use of ITNs [73,74]. To address occupational disparities in ITN ownership and use, tailored health education interventions such as outreach programs that engage farmers in their local settings should be considered. Moreover, providing subsidized ITNs and integrating ITN distribution into broader community health programs can help to ensure equitable access and usage across all occupational sectors [75,76].

Socio-economic status was associated positively with ITNs ownership and use in this study; higher rates observed among individuals from households with higher SES and declined progressively with decreasing wealth index. These findings correlate with those reported elsewhere in sub-Saharan Africa [76,77]. This is likely influenced by financial constraints, as individuals from lower- and middle-income households may not afford ITNs [19,78,79]. There is a need for the government to consider increasing ITN subsidies through both private and public ITN distribution channels such as initiating and implementing special voucher/coupon systems for financially disadvantaged groups to facilitate their ITNs access [45,80,81].

Limitations to this study include the design of the study which was a cross-sectional survey with participants enrolled based on their willingness. This might have introduced selection bias because participants were not selected randomly. Self-reporting data on ITN ownership and use may also be influenced by recall bias or social appeal leading to under or overestimation of actual practices. The study assessed participants’ perceptions of ITNs and assumed that their knowledge on ITNs usage correlated with their education level, an assumption that may not accurately reflect the actual relationship. However, this study offers valuable insights on ITNs access and usage as a core malaria prevention intervention in rural areas. Also, the consistency of these findings with those from previous studies conducted in Tanzania and elsewhere suggests that the degree of selection bias may have been minimal.

## Conclusion

ITN ownership and usage were relatively high among rural community members in the five districts. However, both remained below the 80% target projected by NMCP for 2025. Higher ownership and usage were observed among females, children under five, individuals with higher education levels, and those from households with higher SES. The findings from this study highlight the need for NMCP and its partner to address disparities that hinder equitable ITN access within the current distribution channels to promote universal coverage and consistent use of bed nets across all population groups in Tanzania.

## List of abbreviations

ANC: Antenatal care
BCC: Behaviour change and communication
CIs: Confidence intervals
CSS: Cross-sectional survey
IDs: Identification numbers
IRS: Indoor residual spraying
ITN: Insecticide-treated bed net (plural: ITNs)
IQR: Inter-quantile range
LSM: Larval source management
NIMR: National Institute for Medical Research
MSMT: Molecular Surveillance of Malaria in Tanzania
NMCP: National Malaria Control Programme
ODK: Open Data Kit
PCA: Principal component analysis
SES: Socio-economic status
SNP: School Net Program
TNVS: Tanzania National Voucher Scheme
WHO: World Health Organization

## Declarations

### Ethical approval and consent to participate

This research work was part of the MSMT project with ethical clearance number Ref. NIMR/HQ/R.8a/Vol.1X/35798 dated 16th December 2020, whose protocol was reviewed and approved by the Medical Research Coordinating Committee of National Institute for Medical Research (NIMR). Permission to conduct the study was obtained from the President’s Office, Regional Administration and Local Government (PO-RALG), health authorities of regional and the districts as well as village leaders. Informed consent/assent were sought and obtained from each participant or parents/legal guardians of children before participating in the survey. Permission to publish this paper was sought and obtained from the Director General of NIMR. **Consent for publication**

Not applicable

## Availability of data and materials

Data supporting the findings of this study can be obtained from the corresponding author upon reasonable request, with institutional approval from NIMR and signed data transfer agreement between NIMR and the recipient.

## Author contributions

DSI and GAC conceived the study. DSI, CIM, and MDS supervised the implementation of the CSS and the data analysis conducted by GAC, DAP, SSM and DPC, and contributed to data interpretation. GAC drafted the manuscript, and all authors revised and contributed to the final edition. GAC prepared the final manuscript, which was read and approved by all authors **Acknowledgements**

The authors sincerely thank the study participants for their willingness to participate and take part in this study. We also highly appreciate the support provided by village leaders, districts and regional health authorities throughout the study period. Thanks to the dedicated study teams for their commitment in implementing various aspect of the study, they include Ezekiel Malecela, Filbert Francis, Oswald Oscar, Ildephonce Mathias, Gerion Gaudin, Kusa Mchaina, Hussein Semboja, Sharifa Hassan, Salome Simba, Hatibu Athumani, Ambele Lyatinga, Honest Munishi, Anael Derrick Kimaro, Ally Idrissa, and Amina Ibrahim. We also extend our appreciations to the finance, administrative, and logistics teams at NIMR; Christopher Masaka, Millen Meena, Beatrice Mwampeta, Neema Manumbu, Arison Ekoni, Sadiki Yusuph, John Fundi, Fred Mashanda, Amir Tununu, and Andrew Kimboi for their dedicated support. We greatly appreciate the support from the management of NIMR, NMCP, and PO-RALG, which was critical to the success of this study. The technical and logistical support provided by Brown University, the University of North Carolina at Chapel Hill, the CDC Foundation, and the Gates Foundation team is highly appreciated.

